# Antibody Responses to SARS-Cov-2 among Health Care Workers of a Tertiary Hospital in North-Eastern, Tanzania

**DOI:** 10.1101/2024.04.09.24305582

**Authors:** Pendo M Ibrahim, Felix Anthony, Happiness Mshana, Kevin Rwegoshola, Hadija Semvua, Jaffu Chilongola

## Abstract

**Background:** Health Care Workers (HCWs) have been playing crucial role in treating patient with COVID-19. They have a higher occupational risk of contracting the disease than the general population, and a greater chance of them transmitting the disease to vulnerable patients under their care. Given their scarcity and low COVID-19 vaccine acceptance in Africa, it is essential that HCWs are seroprotected and their exposure to COVID-19 minimized. This study was therefore designed to determine IgG antibody response to SARS-CoV-2 among HCWs in North Eastern, Tanzania.

**Methodology:** This was a cross-sectional study carried out among 273 HCWs at Kilimanjaro Christian Medical Centre (KCMC), a tertiary, zonal referral hospital in Tanzania’s North Eastern region. Stratified sampling was used to select study participants. Data were obtained from each consenting participant using a validated questionnaire. Blood samples were collected for SARS-CoV-2 IgG antibodies quantification by using an indirect ELISA test. RedCap software was used to enter and manage data. Statistical analysis was done by using STATA statistical software version 15 and GraphPad Prism v 9.0. A p-value of < 0.05 was considered the cut-off for statistical significance.

**Results:** Among 273 HCWS only 37.9 % reported to have received COVID-19 vaccine. Except for one person, all of the participants had SARS-CoV-2 IgG antibody concentrations that were positive, with 64.5% of them having strong seropositivity. Female gender, allied health professionals, active smoking, COVID-19 patient interactions, COVID-19 vaccination receptivity, and adherence to recommended hand hygiene were found to be significant predictors of variation of median SARS-CoV-2 antibody concentration. The usage of personal protective equipment, history of previously testing PCR positive for COVID-19, the number of COVID-19 patient exposure and age were found to cause no significant variation in median antibody concentration among participants.

**Conclusions:** This study reports a high seroprevalence of SARS-CoV-2 antibodies among healthcare workers in Kilimanjaro Christian Medical Centre. This suggests that HCWs have significant exposure to SARS-CoV-2 despite the low rate of vaccination acceptance even among HCWs. We recommend a strengthened Infectious Prevention and Control (IPC) in hospitals through provision of technical leadership and coordination according to WHO guidelines. We also recommend continued conduction of seroprevalence studies to estimate the magnitude and trends of SARS-CoV-2 infections in different populations in Tanzania. A better understanding of the past, current, and future transmission patterns of infectious pathogens is critical for preparedness and response planning, and to inform the optimal implementation of existing and novel interventions under the current and changing climate.

## Introduction

The novel corona virus disease 2019 (COVID-19), is caused by the Severe Acute Respiratory Syndrome Coronavirus 2 (SARS-CoV-2)(1,2). COVID-19 has been a large-scale global threat since 2019 with high global morbidity and mortality rates associated with economic disabilities, and social disruptions (3–5). WHO had reported a total of 115,500 deaths of HCWs in the world due to COVID-19 based on population estimates (6). COVID-19 is affects primarily the respiratory system with potential effects in other body organs (1,7). In Tanzania, the first case of COVID-19 was reported in 2020 indicating the virus global spread and its impact (8). Health care personnel have been on the front lines in taking care of COVID-19 patients, thus exposed to a higher occupational risk of contracting the disease than the general population. To protect this vulnerable group, WHO had implemented several initiatives, including making COVID-19 vaccination a priority for HCWs. Nevertheless, there has been a considerably low COVID-19 vaccine acceptance in Africa due to concerns regarding safety and efficacy issues, and associated side effects of the vaccines(9).

Immune responses to SARS-CoV-2 are directed to the four main structural proteins of the virus which are Spike (S), Envelope (E), Membrane (M), and Nucleocapsid (N) proteins (10). A specific humoral immune response against N and S protein has been reported and tend to persist in individuals (11,12). Immune responses to these proteins could be a result of either natural immunity from infection or vaccination (13). However, It has been also been reported that presence of neutralizing antibodies against these proteins correlates with the protection against future SARS-CoV-2 infection (14–16).

There is limited information regarding SARS-Cov-2 immunity among HCWS in Africa. Since antibody response is an acceptable proxy indicator of exposure to an infectious agent, (17), monitoring SARS-CoV-2 antibody response provides important information regarding the burden of exposure to SARS-CoV-2 among the higher risk group of HCWs is important. A better understanding of the past, current, and future transmission patterns of infectious pathogens including emerging and re-emerging infections is critical for preparedness and response planning, and to inform the optimal implementation of existing and novel interventions under the current and changing climate. The current study was designed to assess the seroprevalence of SARS-CoV-2 IgG antibodies among HCWs with different demographics in North-Eastern, Tanzania.

## METHODOLOGY

### Study setting and design

This was a cross sectional study, conducted from September to November, 2022. It was conducted in Kilimanjaro Christian Medical Centre, one of the four tertiary, zonal referral hospitals in Tanzania. It was purposively selected not only because it was a designed national center for managing COVID-19 cases during the pandemic but also its location in North-Eastern region of Tanzania. Kilimanjaro and Arusha are known for being the safari capitals of Tanzania, and popular stopovers for adventurers who are preparing for a Kilimanjaro trek. This makes Kilimanjaro region a vulnerable to cross border transmission of infectious diseases including SARS-CoV-2.

### Study population

This study involved health care workers (HCWs) working at KCMC during the study period. Any person employed or volunteering in this setting was selected based on the definition of a HCW by WHO (18). If the selected HCWs did not consent to participate or donate a blood sample, then they were considered as ineligible for the study and thus excluded.

### Sample Size and Sampling Technique

As there was no prior data on the prevalence of SARS-COV-2 antibodies among HCWs during the design of the study and in order to have sufficient sample size, an estimate of 50% as seroprevalence of SARS COV-2 antibodies among HCWs in Tanzania was considered. Using the formula by (19) and a desired precision of 0.05, and a confidence level of 0.95, a minimum sample size of 257 participants were required. However, to increase the power of analysis, 273 subjects were recruited in this study. To ensure fair healthcare workers’ representation, the population of healthcare workers in KCMC hospital was divided into 13 strata. These strata were represented by different hospital departments. Both inpatient and outpatient healthcare workers were selected from each stratum. Given the busy schedules and responsibilities of these healthcare workers, it was difficult to recruit them systematically for the study in their strata; therefore, a convenience sample of no more than 38 healthcare employees from each stratum was selected.

### Data Collection procedures

Healthcare workers who consented to participate in this study were interviewed by using the study questionnaire embedded in Redcap Software installed on an Android tablet. This was a validated tool by WHO Regional Office for Africa (AFRO) to be used for healthcare workers (20) . Because it was a guidance for SARS-COV-2 antibody screening among HCWs for cohort studies, only questions used for participant enrollment were asked in this study. This adapted questionnaire included socio demographic and clinical characteristics, information about COVID-19 vaccination history, and COVID-19 illness, occupation and community-related behaviour during the pandemic.

### Sample Collection

From each study participant a total of 2 mls of blood sample through vein puncture were collected under aseptic condition. Samples were stored in a cooler box (maintained at 4-8°C) in the field for a maximum of 3 hours before these samples get transferred to the Biotechnology Laboratory at Kilimanjaro Christian Research Institute for serum extraction. The samples that were collected had instantly their serum extracted upon arrival at Biotechnology Laboratory at Kilimanjaro Christian Research Institute. For serum extraction, samples were allowed to clot then they were centrifuged at 1000 g for 15 minutes. After that, the serum was collected and kept frozen at negative 20°C.

### Detection of SARS-COV-2 Antibodies

IgG antibodies against SARS-CoV-2 were detected by using Generic Assays (GA) Enzyme-Linked Immuno-Sorbent assay (ELISA) for SARS-CoV-2 IgG Screening kits (MedipanGmbHGA Generic Assays GmbH, Ludwig-Erhard-Ring 3, 15827 Blankenfelde-Mahlow OT Dahlewitz, Germany). This indirect ELISA kit was a two-stage that focuses on the Spike and Nucleocapsid antigen of the SARS-CoV-2 virus detection. .The reported sensitivity and specificity of these GA ELISA tests are > 98% (21).

### Statistical analysis

STATA statistical software version 15 was used to do all statistical tests. Hence, all data from the created spreadsheet was imported to STATA. Descriptive statistics were employed to summarize the study participant’s baseline socio-demographic, clinical, COVID-19 exposure history as well as the seroprevalence of antibodies against SARS-CoV-2. After verifying that SARS-CoV-2 IgG concentration among HCWs is not normally distributed (p =0.00132 by Shapiro Wilk test), non-parametric tests were performed to compare the significant difference between the exposure variables and median SARS-CoV-2 IgG concentration. The Mann–Whitney test was used for the comparison of antibody concentrations of two independent groups. The Kruskal–Wallis test was used for the comparison of more than two groups. A p-value of 0.05 was regarded as significant for the found associations

### Ethical Considerations

Ethical clearance to carry out this study was obtained from the College Research Ethical Committee (CREC) of Kilimanjaro Christian Medical University College (KCMUCo), ethical clearance number PG61/2022. Permission from the participant hospital administration was sought after the proposal was submitted and accepted by the ethical committee. To conceal participants’ identities, the questionnaire and blood samples were labelled using numbers and letters.

## RESULTS

### Response rate

A total of 273 of the 279 participants in this study had results on their serum SARS-CoV-2 IgG concentration, resulting in a rate of response of 97.8%.

### Demographic and Clinical-exposure Characteristics of the Study Participants

Among the 273 participants, half of the participants were under 32 years old, with a median age of 32 (IQR: 26-44) and a male predominance of 60.4% among the total number of participants. The majority of study participants were nurses (40.5 %) and had a normal BMI (41.3%). Less than half of the study participants received the COVID-19 vaccine, and only 8.8% reported being tested PCR positive for COVID-19 in the past. The vast majority of participants (94.1%) stated that they had never smoked, Table 1.

**Table 1:** Social demographic and clinical characteristics of the study participants (N=273)

| Variable | Frequency | Percentage |
| --- | --- | --- |
| **Sex** |  |  |
| Male | 165 | 60.4 |
| Female | 108 | 39.6 |
| **Age (in Years) \*(n=272)** |  |  |
| ≤ 32 years | 142 | 52.2 |
| > 32 | 130 | 47.8 |
| *Median (IQR)* | 32 (26-44) |  |
| **Cadre\*(n=268)** |  |  |
| Medical doctor | 78 | 29.0 |
| Nurse | 109 | 40.5 |
| Allied health professionals | 58 | 21.6 |
| Support staff | 23 | 8.9 |
| **BMI\*(n=267)** |  |  |
| Underweight | 6 | 2.3 |
| Normal | 109 | 40.8 |
| Overweight | 83 | 31.1 |
| Obesity | 69 | 25.8 |
| *Median (IQR)* | 26.4(22.8-30.1) |  |
| **Smoking status** |  |  |
| Stopped >1 year ago | 8 | 2.9 |
| Never smoked | 257 | 94.1 |
| Currently smoke | 8 | 3.0 |
| **Alcohol consumption** |  |  |
| Stopped >1 year ago | 17 | 6.2 |
| Never took alcohol | 158 | 57.8 |
| Currently take alcohol | 98 | 36.0 |
| **Taking regular medication** |  |  |
| No | 233 | 85.3 |
| Yes | 40 | 14.7 |
| **Tested PCR Positive for COVID-19\*(n=272)** |  |  |
| No | 248 | 91.2 |
| Yes | 24 | 8.8 |
| **Received COVID-19 vaccine\*(n=272)** |  |  |
| No | 169 | 62.1 |
| Yes | 103 | 37.9 |

\* Indicates some missing values in respective variable

### Occupation and Community Related Behaviour Factors during the Pandemic

Only 38.5% of the study participants wore mask at indoor setting outside their homes. A large proportion of participants (56.6%) practice good hand hygiene always as recommended, 38.9% follow IPC standard precautions when in contact with any patients, and fewer than half (42.4%) always wear PPE according to the risk assessment. Half of the study participant lived in a household size of 3-5 people and 39.5% used public transportation more than nine times a day, Table 2.

**Table 2:** Behavioural characteristics of study participants (N=273)

| Variable | Frequency | Percentage |
| --- | --- | --- |
| **Household size\*(n=272)** |  |  |
| 1-2 people | 90 | 33.1 |
| 3-5 people | 136 | 50.0 |
| 6-8 people | 37 | 13.6 |
| 9+ | 9 | 3.3 |
| **Public transport** |  |  |
| None | 75 | 27.5 |
| 1-2/day | 68 | 24.9 |
| 3-5/day | 19 | 7.0 |
| 6-8/day | 3 | 1.1 |
| 9+/day | 108 | 39.5 |
| **Stayed at least 2 meters from other people in indoor space\*(n=273)** |  |  |
| Always | 42 | 15.4 |
| Did not go indoor location | 31 | 11.4 |
| Never | 31 | 11.4 |
| Often | 28 | 10.2 |
| Rarely | 56 | 20.5 |
| Sometimes | 85 | 31.1 |
| **Hand hygiene practice\*(n=265)** |  |  |
| Always as recommended | 150 | 56.6 |
| Most of the time | 104 | 39.3 |
| Never | 3 | 1.1 |
| Occasionally | 8 | 3.0 |
| **IPC standards\*(n=257)** |  |  |
| Always | 100 | 38.9 |
| I don't know what IPC standard precaution means | 22 | 8.6 |
| Most of the time | 97 | 37.7 |
| Never | 2 | 0.8 |
| Occasionally | 28 | 10.9 |
| Rarely | 8 | 3.1 |
| **Wearing PPE as recommended\*(n=264)** |  |  |
| Always | 112 | 42.4 |
| Most of the time | 102 | 38.6 |
| Never | 8 | 3.1 |
| Occasionally | 33 | 12.5 |
| Rarely | 9 | 3.4 |
| **Interactions with COVID-19 Patients\*(n=264)** |  |  |
| No | 110 | 41.7 |
| Yes | 154 | 58.3 |
| **Exposure to COVID-19 Patients\*(n=247)** |  |  |
| 1-10 Patients | 169 | 68.4 |
| 11-50 | 41 | 16.6 |
| 51-100 | 18 | 7.3 |
| 101-500 | 17 | 6.9 |
| > 500 | 2 | 0.8 |

*\*Indicates some missing values in respective variable*

### Seroprevalence of SARS-CoV-2 IgG Antibody Concentration among the Study Participants

Except for one person, all of the participants showed SARS-CoV-2 IgG antibody concentrations that were positive, with 64.5% of them having strong seropositivity, Figure 1. While comparing people who had received the COVID-19 vaccine and those who hadn’t, it was shown that the majority of the vaccinated individuals had strong seropositivity, Figures 2 and 3.

**Figure 1:**
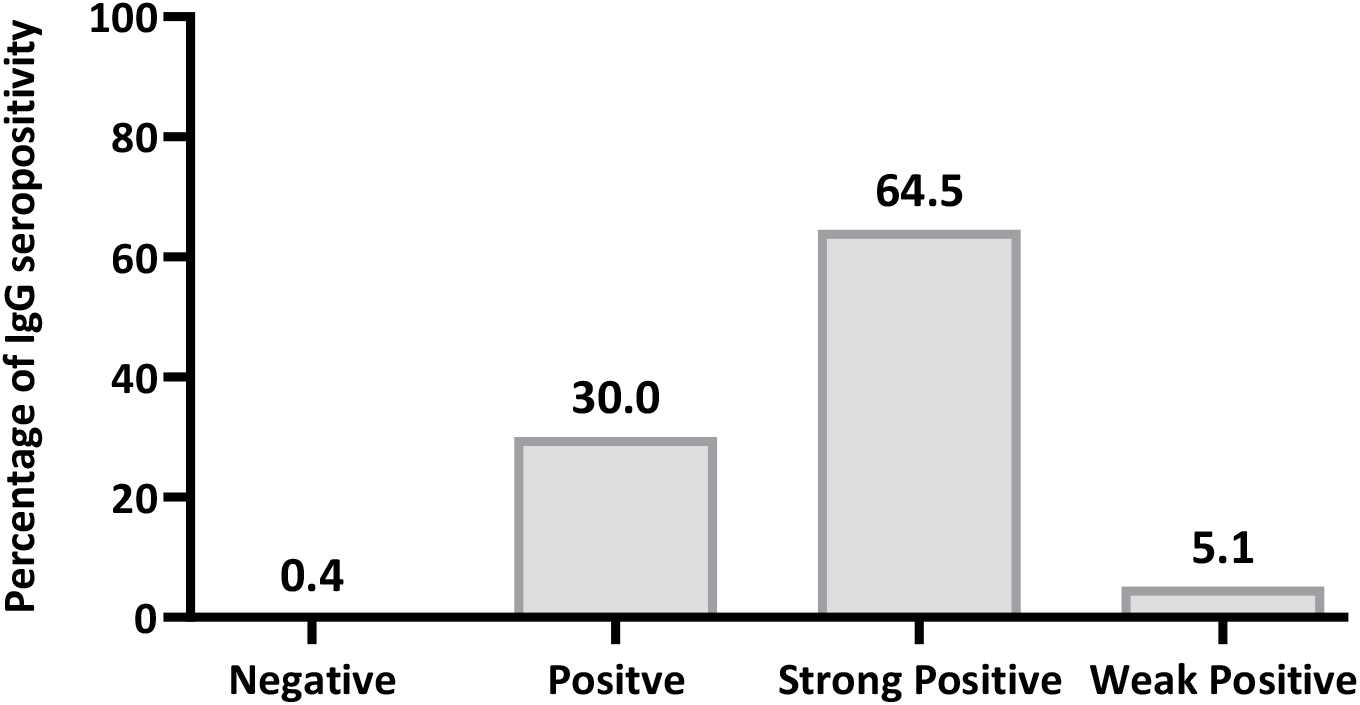
Seroprevalence of SARS-CoV-2 IgG antibody concentrations among the study participants (N=273)

**Figure 2:**
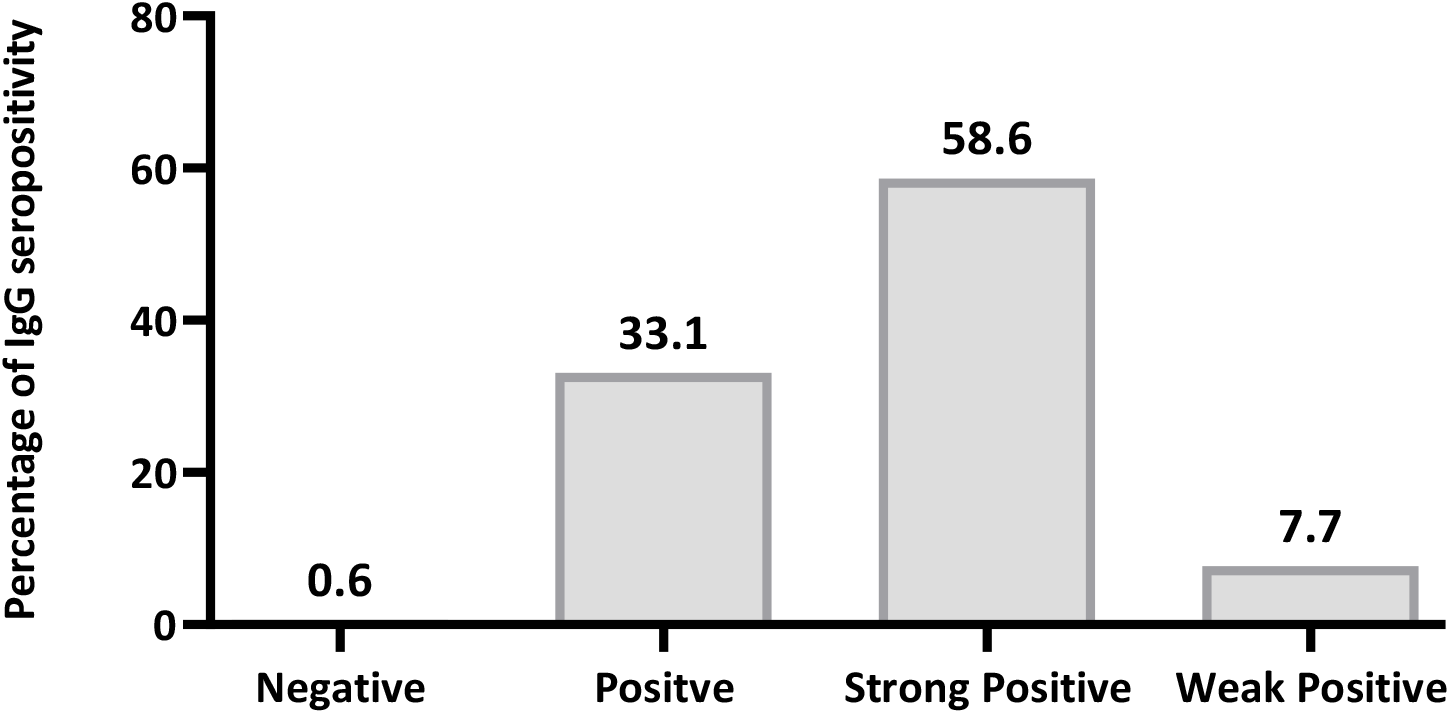
Seroprevalence of SARS-CoV-2 IgG antibody concentrations among non-vaccinated participants (N=169)

**Figure 3:**
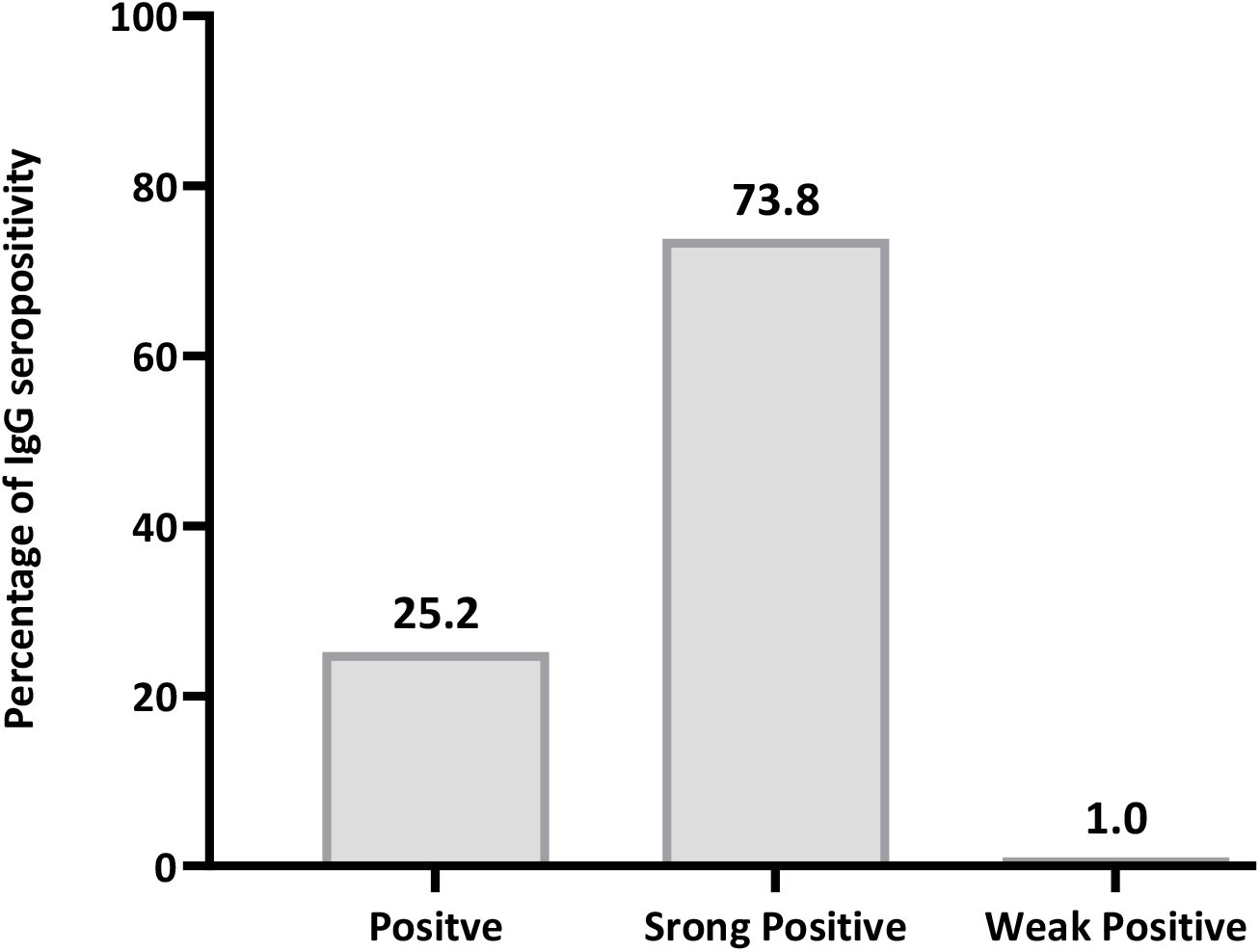
Seroprevalence of SARS-CoV-2 IgG antibody concentrations among vaccinated participants (N=103)

### Socio-Demographic, Clinical, and Behavioural Characteristics Associated with Variation in Median SARS-CoV-2 IgG Concentration among Study Participants

Sex, BMI, smoking status, adherence to recommended hand hygiene, cadre, and interaction with COVID-19 patients are variables that were found to significantly affect the median IgG concentration. IgG median concentration was significantly higher in females compared to males. It was found that those with obesity had significantly greater median concentrations than individuals with other BMI categories. Those who had never smoked had significantly higher SARS-CoV-2 IgG median concentrations than current smokers. Individuals, who followed recommended hand hygiene were found to significantly have a higher median concentration. Moreover, median concentrations were significantly greater in who interacted with COVID-19 patients. Interestingly, allied health proffessionals were found to have a significantly higher median concentration comparing to other health care workers. Other factors were assessed but found to not significantly causing differences in median SARSCOV 2 IgG concentration among participants, **Figure 4**.

**Figure 4:**
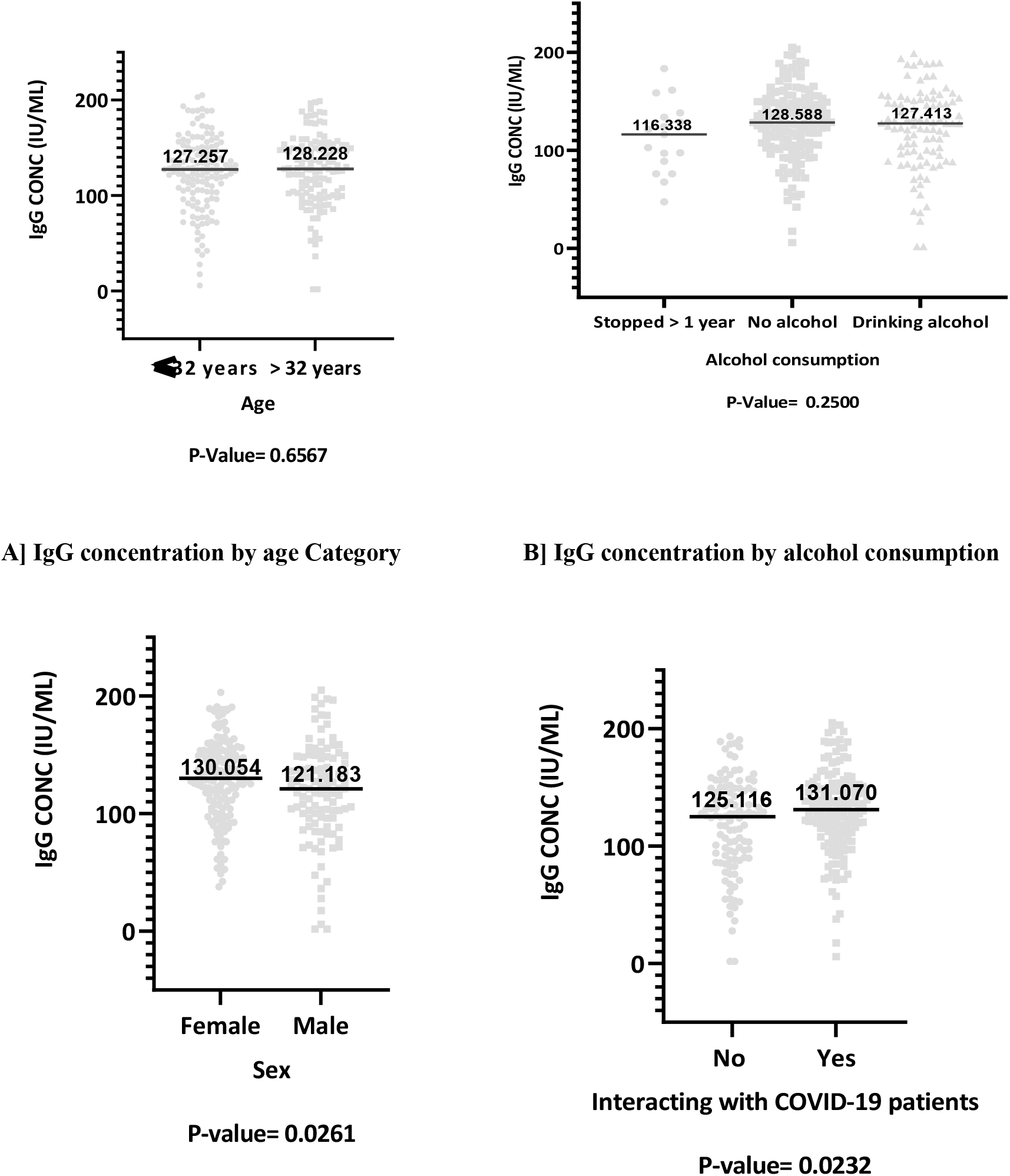
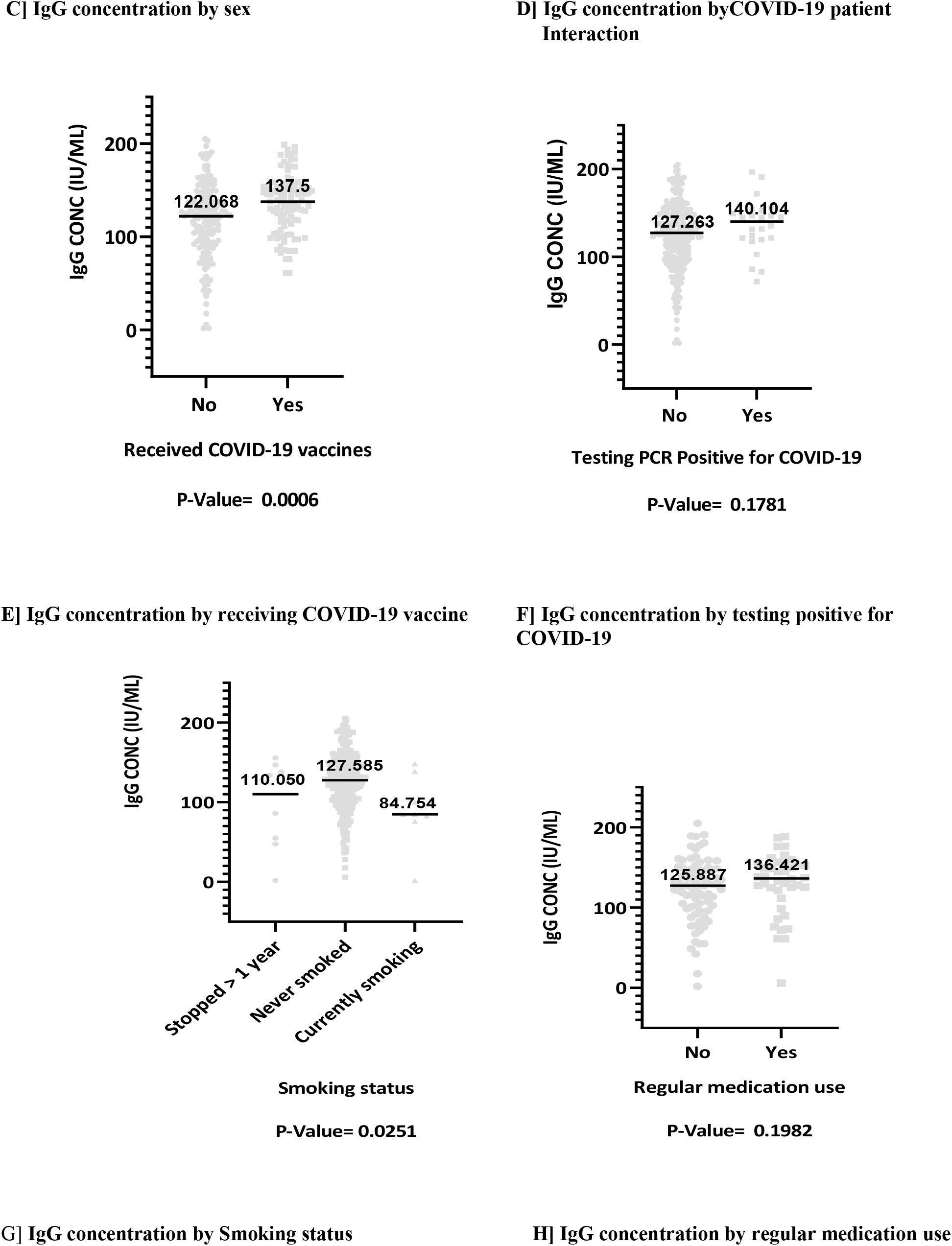
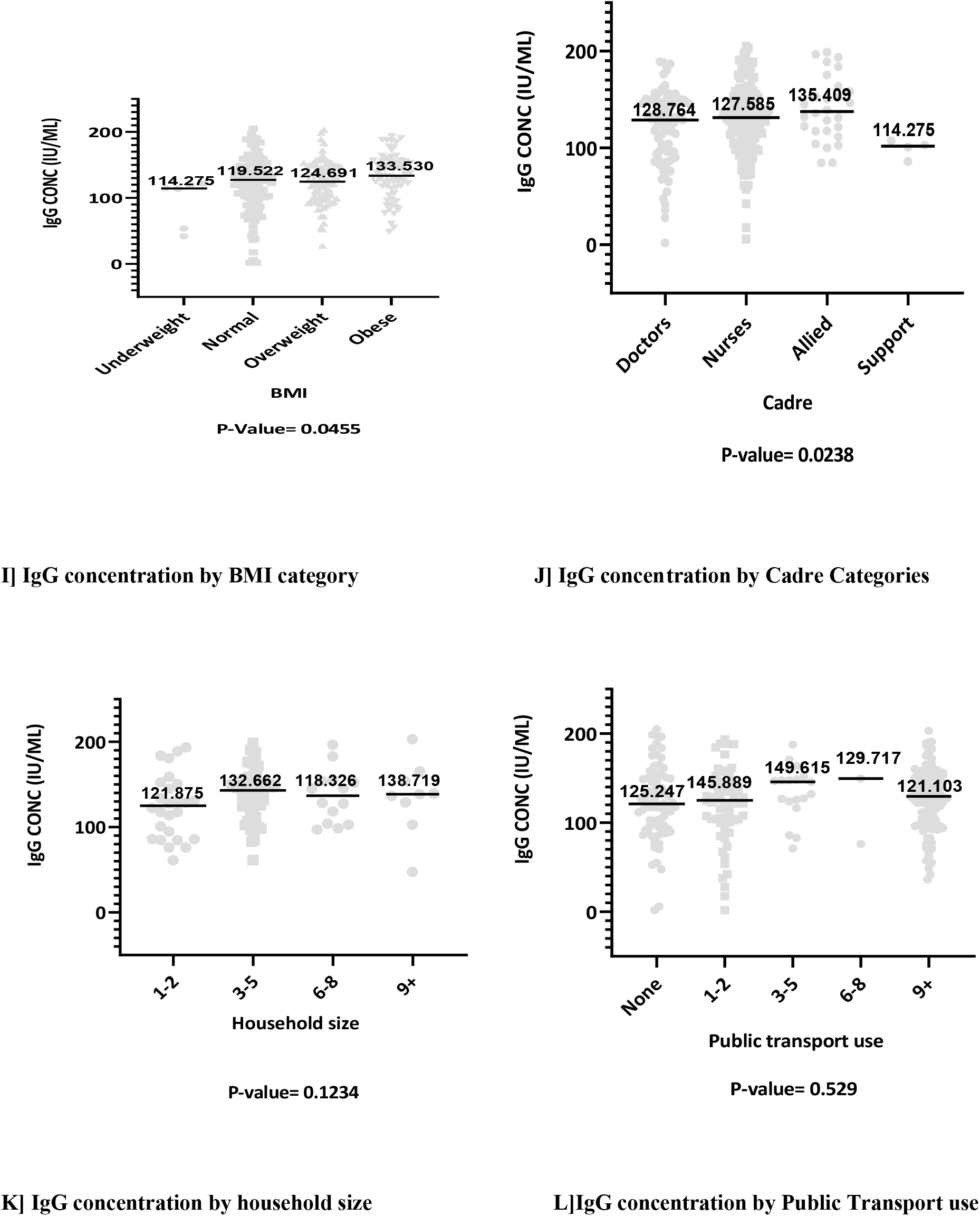
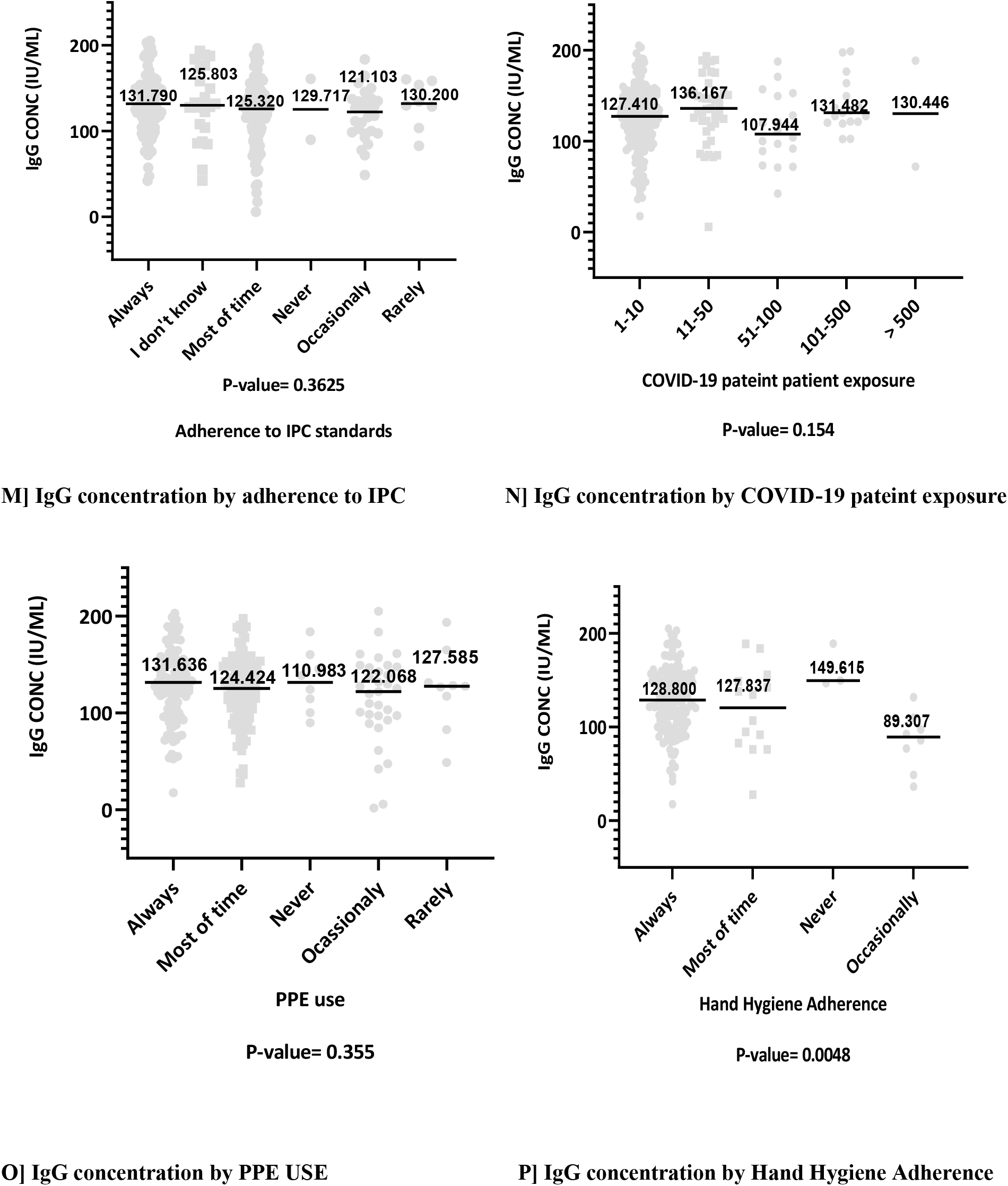
The median difference in IgG concentration among participants in different exposure groups.

## DISCUSSION

This study aimed at determining the IgG antibody response to SARS-CoV-2 among HCWs in our institution. The finding revealed a remarkably seroprevalence of 99.6% among the sampled HCWs. Females, allied health professionals, obese people, HCWs who adhered to recommended hand hygiene practices, and those who interacted with COVID-19 patients more frequently had significantly higher median SARS-CoV-2 antibody concentrations. Other factors that were assessed did not reveal any significant variation in Median SARS-CoV-2 IgG concentration.

This higher seroprevalence of SARS-CoV-2 antibodies among HCWs indicates the high level of virus exposure in this population, and an existed risk of infection within the hospital. These results are also consistent with other studies that revealed high seroprevalence of SARS-CoV-2 antibodies among HCWs (22,23). However, this seroprevalence is higher than that reported in other East African countries (24–26). The level of COVID-19 pandemic precautions that was initially put in our country can explain this huge discrepancy with other East African countries.

Contrary to the expectation, this study did not find any significant difference in antibody concentration between healthcare workers considering their previous history of testing PCR positive for COVID-19. This is contradicting finding to other previous studies that indicated that previous SARS-CoV-2 infection leads to higher SARS-CoV-2 IgG antibodies concentrations (27–30). Several factors may have influenced these results. It might be due to the low percentage of healthcare workers who tested positive in this study. Also, the significant decline of SARS-CoV-2 antibody levels after infection may additionally explain this results (31). Hence, a time interval for antibody monitoring should be found so as to determine how long SARS-CoV-2 antibodies does lasts.

Regarding various cadres, this study found that allied health professional had a higher SARS-CoV-2 median concentration compared to other HCWs. This indicate that there was an increased risk of SARS-CoV-2 exposure among allied HCWS compared medical doctors and nursing cadres. Results are concordant with one previous observational study, which had shown increasing odds off seropositivity in allied health professionals compared to medical doctors (32). The reasons behind this finding are necessary to be explored in order to protect these allied HCWs from the risk of acquiring communicable diseases in their work setting

Another important finding was a higher median antibody concentration among healthcare workers who adhere to recommended hand hygiene during the pandemic. The finding of our study does not support other studies that found no association between self-reported adherence to hand hygiene and SARS-CoV-2 antibody positivity among HCWs (33). Hand hygiene is an important element of infection prevention practices in the hospital and reflects behavior, attitudes and beliefs (34). It may be hypothesized that HCWs who adhered to recommended hand hygiene were also more likely to receive COVID-19 vaccine. However, this hypothesis was not explored in this study.

Study finding has revealed that HCWs who interacted with COVID-19 patient had significantly higher median concentration. It is important to note that the number of COVID-19 patient the HCW is exposed to, does not predict seroconversion as per our study findings. Therefore, it confirms that COVID-19 patient exposure only is a significant factor for detecting SARS-CoV-2 antibodies among HCWs. These results match those observed in earlier studies that demonstrated that regular interaction with COVID-19 patients increases one’s risk of contracting SARS-CoV-2 (27,35–39). It can be hypothesized that relying solely in number of COVID-19 patient in hospitals led to inadequate HCWs protection.

According to our findings, females had higher median concentrations of SARS-CoV-2 antibodies than males. This supports the theory that after disease exposure, females have higher antibody production compared to males because male androgens suppress the immune response (40). Contrary to our findings, several other studies have found that male HCWs have higher SARS-CoV-2 antibodies than female HCWs, inferring to behavioural differences (25,27,37,41,42). During the pandemic, male had a worse clinical outcomes and mortality (Kopel *et al*., 2020; O’Brien, Du and Peng, 2020). The argument can be that the immune response of female played a major role in Clinical outcome of COVID-19 rather than behavioural differences with males.

The findings from this study indicate that individuals who currently smoke had a lower antibody response to SARS-CoV-2. This may be due to the fact that smoking increases the clearance of circulating antibodies by enhancing the production of monocytes and macrophages (43). It can also can be evidenced by the reduction of antibody titers after COVID-19 vaccination in individuals who smoke (44). However, some other studies have found no association between smoking and SARS-CoV-2 antibody concentrations (45,46). The study methodology and demographic characteristics between studies may explain this finding variations between studies.

### 5.1 Conclusion

This study has revealed a high prevalence of SARS-CoV-2 IgG antibodies among HCWs in our setting. These results reveal that most HCWs are exposed to SARS-CoV-2 infection and thus probably more seroprotected.

### 5.2 Study limitations and strengths of the study

This study has successfully demonstrated its purpose, but the scope of this study was limited as such, retrospective assessment of self-reported exposures may be subject to recall bias. This study was conducted in one centre, a tertiary hospital where exposure to the SARS-CoV-2 virus cannot be underestimated. Therefore, a caution must be applied in a nationwide result generalization. Notwithstanding these limitations, this is the first study surveying the seroprevalence of SARS-CoV-2 IgG antibodies among healthcare workers in Tanzania.

### 5.3 Recommendation

This research has thrown up many questions in need of further investigation. More broadly, future research should focus in large cohort to determine the long-term implication of SARS-CoV-2 antibodies such as probability of reinfection and the outcome of vaccination uptake among HCWS in our setting. A reasonable approach tackle HCWs disease exposures is to implement strict preventive measures for communicable diseases in society to reduce the burden in hospitals.

## Data Availability

All data produced in the present study are available upon reasonable request to the authors.

## References

1. Harapan H, Itoh N, Yufika A, Winardi W, Keam S, Te H, et al. Coronavirus disease 2019 (COVID-19): A literature review. Journal of Infection and Public Health. 2020 May 1;13(5):667–73.

2. Zhu N, Zhang D, Wang W, Li X, Yang B, Song J, et al. A Novel Coronavirus from Patients with Pneumonia in China, 2019. N Engl J Med. 2020 Feb 20;382(8):727–33.

3. Poudel K, Subedi P. Impact of COVID-19 pandemic on socioeconomic and mental health aspects in Nepal. Int J Soc Psychiatry. 2020 Dec;66(8):748–55.

4. Wang C, Wang D, Abbas J, Duan K, Mubeen R. Global Financial Crisis, Smart Lockdown Strategies, and the COVID-19 Spillover Impacts: A Global Perspective Implications From Southeast Asia. Frontiers in Psychiatry [Internet].2021 [cited 2023 May 19];12. Available from: https://www.frontiersin.org/articles/10.3389/fpsyt.2021.643783

5. Lekagul A, Chattong A, Rueangsom P, Waleewong O, Tangcharoensathien V. Multi-dimensional impacts of Coronavirus disease 2019 pandemic on Sustainable Development Goal achievement. Globalization and Health. 2022 Jun 27;18(1):65.

6. World Health Organization. The impact of COVID-19 on health and care workers: a closer look at deaths [Internet]. Geneva: World Health Organization; 2021. Available from: https://apps.who.int/iris/handle/10665/345300

7. Mehta OP, Bhandari P, Raut A, Kacimi SEO, Huy NT. Coronavirus Disease (COVID-19): Comprehensive Review of Clinical Presentation. Frontiers in Public Health [Internet]. 2021 [cited 2023 May 19];8. Available from: https://www.frontiersin.org/articles/10.3389/fpubh.2020.582932

8. Tarimo CS, Wu J. The first confirmed case of COVID-19 in Tanzania: recommendations based on lesson learned from China. Tropical Medicine and Health. 2020 Apr 26;48(1):25.

9. Ackah M, Ameyaw L, Gazali Salifu M, Afi Asubonteng DP, Osei Yeboah C, Narkotey Annor E, et al. COVID-19 vaccine acceptance among health care workers in Africa: A systematic review and meta-analysis. PLoS One. 2022;17(5):e0268711.

10. Satarker S, Nampoothiri M. Structural Proteins in Severe Acute Respiratory Syndrome Coronavirus-2. Arch Med Res. 2020 Aug;51(6):482–91.

11. Smits VAJ, Hernández-Carralero E, Paz-Cabrera MC, Cabrera E, Hernández-Reyes Y, Hernández-Fernaud JR, et al. The Nucleocapsid protein triggers the main humoral immune response in COVID-19 patients. Biochem Biophys Res Commun. 2021 Mar 5;543:45–9.

12. Wu J, Liang B, Chen C, Wang H, Fang Y, Shen S, et al. SARS-CoV-2 infection induces sustained humoral immune responses in convalescent patients following symptomatic COVID-19. Nat Commun. 2021 Mar 22;12:1813.

13. Suhandynata RT, Bevins NJ, Tran JT, Huang D, Hoffman MA, Lund K, et al. SARS-CoV-2 Serology Status Detected by Commercialized Platforms Distinguishes Previous Infection and Vaccination Adaptive Immune Responses. The journal of applied laboratory medicine. 2021;6(5):1109–22.

14. Alsoussi WB, Turner JS, Case JB, Zhao H, Schmitz AJ, Zhou JQ, et al. A Potently Neutralizing Antibody Protects Mice against SARS-CoV-2 Infection. The Journal of Immunology. 2020 Aug 15;205(4):915– 22.

15. Deng W, Bao L, Liu J, Xiao C, Liu J, Xue J, et al. Primary exposure to SARS-CoV-2 protects against reinfection in rhesus macaques. Science. 2020 Jul 2;eabc5343.

16. Earle KA, Ambrosino DM, Fiore-Gartland A, Goldblatt D, Gilbert PB, Siber GR, et al. Evidence for antibody as a protective correlate for COVID-19 vaccines. Vaccine. 2021 Jul 22;39(32):4423–8.

17. Zheng J, Deng Y, Zhao Z, Mao B, Lu M, Lin Y, et al. Characterization of SARS-CoV-2-specific humoral immunity and its potential applications and therapeutic prospects. Cell Mol Immunol. 2022 Feb;19(2):150–7.

18. Global Health Workforce statistics database [Internet]. [cited 2022 Aug 6]. Available from: https://www.who.int/data/gho/data/themes/topics/health-workforce

19. Sample size calculation in medical studies - PMC [Internet]. [cited 2023 Mar 21]. Available from: https://www.ncbi.nlm.nih.gov/pmc/articles/PMC4017493/

20. WHO | Regional Office for Africa [Internet]. 2023 [cited 2023 May 17]. Guidance Document Cohort study to measure COVID-19 vaccine effectiveness among health workers. Available from: https://www.afro.who.int/publications/guidance-document-cohort-study-measure-covid-19-vaccine-effectiveness-among-health

21. Deshpande K, Pt U, Kaduskar O, Vijay N, Rakhe A, Vidhate S, et al. Performance assessment of seven SARS-CoV-2 IgG enzyme-linked immunosorbent assays. J Med Virol. 2021 Dec;93(12):6696–702.

22. Murhekar M, Bhatnagar T, Selvaraju S, Rade K, Saravanakumar V, Vivian Thangaraj J, et al. Prevalence of SARS-CoV-2 infection in India: Findings from the national serosurvey, May-June 2020. Indian J Med Res. 2020;152(1):48.

23. Hajissa K, Islam MA, Hassan SA, Zaidah AR, Ismail N, Mohamed Z. Seroprevalence of SARS-CoV-2 Antibodies in Africa: A Systematic Review and Meta-Analysis. Int J Environ Res Public Health. 2022 Jun 14;19(12):7257.

24. Etyang AO, Lucinde R, Karanja H, Kalu C, Mugo D, Nyagwange J, et al. Seroprevalence of Antibodies to Severe Acute Respiratory Syndrome Coronavirus 2 Among Healthcare Workers in Kenya. Clin Infect Dis. 2021 Apr 23;74(2):288–93.

25. Gelanew T, Seyoum B, Mulu A, Mihret A, Abebe M, Wassie L, et al. High seroprevalence of anti-SARS-CoV-2 antibodies among Ethiopian healthcare workers. BMC Infect Dis. 2022 Dec;22(1):1–9.

26. Ssuuna C, Galiwango RM, Kankaka EN, Kagaayi J, Ndyanabo A, Kigozi G, et al. Severe Acute Respiratory Syndrome Coronavirus-2 seroprevalence in South-Central Uganda, during 2019–2021. BMC Infectious Diseases. 2022 Feb 21;22(1):174.

27. Galanis P, Vraka I, Fragkou D, Bilali A, Kaitelidou D. Seroprevalence of SARS-CoV-2 antibodies and associated factors in healthcare workers: a systematic review and meta-analysis. Journal of Hospital Infection. 2021 Feb;108:120–34.

28. Napolitano F, Di Giuseppe G, Montemurro MV, Molinari AM, Donnarumma G, Arnese A, et al. Seroprevalence of SARS-CoV-2 Antibodies in Adults and Healthcare Workers in Southern Italy. International Journal of Environmental Research and Public Health. 2021 Jan;18(9):4761.

29. Papasavas P, Olugbile S, Wu U, Robinson K, Roberts AL, O’Sullivan DM, et al. Seroprevalence of SARS-CoV-2 antibodies, associated epidemiological factors and antibody kinetics among healthcare workers in Connecticut. J Hosp Infect. 2021 Aug;114:117–25.

30. Decarreaux D, Pouquet M, Souty C, Vilcu AM, Prévot-Monsacre P, Fourié T, et al. Seroprevalence of SARS-CoV-2 IgG Antibodies and Factors Associated with SARS-CoV-2 IgG Neutralizing Activity among Primary Health Care Workers 6 Months after Vaccination Rollout in France. Viruses. 2022 May;14(5):957.

31. Yang Y, Yang M, Peng Y, Liang Y, Wei J, Xing L, et al. Longitudinal analysis of antibody dynamics in COVID-19 convalescents reveals neutralizing responses up to 16 months after infection. Nat Microbiol. 2022 Mar;7(3):423–33.

32. Kribi S, Touré F, Mendes A, Sanou S, Some A, Aminou AM, et al. Multicountry study of SARS-CoV-2 and associated risk factors among healthcare workers in Côte d’Ivoire, Burkina Faso and South Africa. Transactions of The Royal Society of Tropical Medicine and Hygiene. 2023 Mar 1;117(3):179– 88.

33. Sharma P, Chawla R, Bakshi R, Saxena S, Basu S, Bharti PK, et al. Seroprevalence of antibodies to SARS-CoV-2 and predictors of seropositivity among employees of a teaching hospital in New Delhi, India. PHRP. 2021 Apr 9;12(2):88–95.

34. Mathur P. Hand hygiene: Back to the basics of infection control. Indian J Med Res. 2011 Nov;134(5):611–20.

35. Garcia-Basteiro AL, Moncunill G, Tortajada M, Vidal M, Guinovart C, Jiménez A, et al. Seroprevalence of antibodies against SARS-CoV-2 among health care workers in a large Spanish reference hospital. Nat Commun. 2020 Dec;11(1):3500.

36. Almaghrabi RS, Alsagheir OI, Alquaiz RM, Alhekail OZ, Abaalkhail AM, Alduaij AA, et al. Seroprevalence of SARS-CoV-2 antibodies in healthcare workers at a tertiary care hospital in Riyadh, Saudi Arabia. IJID Reg. 2021 Nov 29;2:51–4.

37. Hossain A, Nasrullah SM, Tasnim Z, Hasan MdK, Hasan MdM. Seroprevalence of SARS-CoV-2 IgG antibodies among health care workers prior to vaccine administration in Europe, the USA and East Asia: A systematic review and meta-analysis. EClinicalMedicine. 2021 Mar 8;33:100770.

38. Ken-Dror G, Wade C, Sharma SS, Irvin-Sellers M, Robin J, Fluck D, et al. SARS-CoV-2 antibody seroprevalence in NHS healthcare workers in a large double-sited UK hospital. Clinical Medicine. 2021 May 1;21(3):e290–4.

39. Sonmezer MC, Erul E, Sahin TK, Rudvan Al I, Cosgun Y, Korukluoglu G, et al. Seroprevalence of SARS-CoV-2 Antibodies and Associated Factors in Healthcare Workers before the Era of Vaccination at a Tertiary Care Hospital in Turkey. Vaccines. 2022 Feb;10(2):258.

40. Sciarra F, Campolo F, Franceschini E, Carlomagno F, Venneri MA. Gender-Specific Impact of Sex Hormones on the Immune System. International Journal of Molecular Sciences. 2023 Jan;24(7):6302.

41. Kayi I, Madran B, Keske Ş, Karanfil Ö, Arribas JR, Pshenichnaya N, et al. The seroprevalence of SARS-CoV-2 antibodies among health care workers before the era of vaccination: a systematic review and meta-analysis. Clinical Microbiology and Infection. 2021 Sep 1;27(9):1242–9.

42. Pagheh AS, Asghari A, Romenjan KA, Mousavi T, Abedi F, Ziaee A, et al. Seroprevalence of SARS-COV-2 antibodies among health-care workers exposed to COVID-19 patients in a large reference hospital, Iran. Iranian Journal of Microbiology. 2022 Apr;14(2):138.

43. Pedersen KM, Çolak Y, Ellervik C, Hasselbalch HC, Bojesen SE, Nordestgaard BG. Smoking and increased white and red blood cells: a mendelian randomization approach in the copenhagen general population study. Arteriosclerosis, thrombosis, and vascular biology. 2019;39(5):965–77.

44. Ferrara P, Ponticelli D, Agüero F, Caci G, Vitale A, Borrelli M, et al. Does smoking have an impact on the immunological response to COVID-19 vaccines? Evidence from the VASCO study and need for further studies. Public Health. 2022 Feb;203:97–9.

45. Grégoire C, Huynen P, Gofflot S, Seidel L, Maes N, Vranken L, et al. Predictive factors for the presence and long-term persistence of SARS-CoV-2 antibodies in healthcare and university workers. Scientific Reports [Internet]. 2022 [cited 2022 Aug 1];12. Available from: https://www.ncbi.nlm.nih.gov/pmc/articles/PMC9191528/

46. Halili R, Bunjaku J, Gashi B, Hoxha T, Kamberi A, Hoti N, et al. Seroprevalence of anti-SARS-CoV-2 antibodies among staff at primary healthcare institutions in Prishtina. BMC Infect Dis. 2022 Dec;22(1):57.

